# Outpatient therapies for COVID-19: How do we choose?

**DOI:** 10.1101/2021.12.17.21268007

**Authors:** Todd C. Lee, Andrew M. Morris, Steven A. Grover, Srinivas Murthy, Emily G. McDonald

## Abstract

**Background:** Several outpatient COVID-19 therapies have reduced hospitalization in randomized controlled trials. The choice of therapy may depend on drug efficacy, toxicity, pricing, availability, and access to administration infrastructure. To facilitate comparative decision making, we evaluated the efficacy of each treatment in clinical trials and then estimated the associated cost per hospitalization prevented.

**Methods:** Wherever possible, we obtained relative risk for hospitalization from published randomized controlled trials. Otherwise, we extracted data from press releases, conference abstracts, government submissions, or preprints. If more than one study was published, the results were meta-analyzed. Using relative risk, we estimated the number needed to treat (NNT), assuming a baseline hospitalization risk of 5%. Drug pricing was based on Canadian formularies, government purchases, or manufacturer estimates.

Administrative and societal costs were not included. Results will be updated online as new studies emerge or final publication numbers become available.

**Results:** At a 5% risk of hospitalization the estimated NNTs were: 87 for colchicine, 80 for fluvoxamine, 72 for inhaled corticosteroids, 24 for nirmatrelvir/ritonavir, 25 for sotrovimab, 24 for remdesivir, 29 for casirivimab/imdevimab, 29 for bamlanivimab/etesevimab and 52 for molnupiravir. Colchicine, fluvoxamine, inhaled corticosteroids, and nirmatrelvir/ritonavir had cost per hospitalization prevented point estimates below the CIHI estimated cost of hospitalization ($23000).

**Interpretation:** Canada is fortunate to have access to several effective outpatient therapies to prevent COVID-19 hospitalization. Given differences in efficacy, toxicity, cost and administration complexities, this assessment serves as one tool to help guide policy makers and clinicians in their treatment selection.

## Introduction

The COVID-19 pandemic has fueled an explosion of scientific inquiry. Since the initial reports of overwhelmed health systems and hospitals, there has been tremendous interest in finding outpatient treatments which could prevent hospitalization among those who are symptomatic and at high risk for clinical deterioration. Initial studies looked at drug repurposing: identifying widely available, inexpensive, and safe medications that could prove effective. Initially, hydroxychloroquine was considered a leading candidate (1); however, interest waned as randomized controlled trial evidence failed to demonstrate superiority over placebo (2,3). Since that time, there have been a number of promising repurposed medications including colchicine (4), inhaled corticosteroids (5), and fluvoxamine (6,7) all of which have shown a relative risk reduction of 20-30% in hospitalization. Novel therapeutics have emerged, such as customized anti-spike protein monoclonal antibody products, which have shown up to a 55-85% relative risk reduction in hospitalization (8–10). However, these therapies are not always widely available, are more challenging to administer, comparatively expensive, and may have reduced efficacy against newer variants. Most recently, repurposed and novel antiviral therapies have attracted attention, with relative risk reductions of 30-85% (11–15). Some governments, including Canada, have proactively purchased these agents based on pre-published data (16,17).

For policy makers and/or healthcare professionals, especially those without ready access to novel therapeutics, the decision might be between supportive care or repurposed drugs. For well-resourced healthcare systems such as our own, the anti-spike monoclonal antibodies, remdesivir, and oral antiviral therapies cost significantly more, are available in relatively limited quantities, and can be more complex to procure and/or administer. Our objective was to systematically quantify the effect sizes of available treatments with respect to preventing hospitalization and then to contextualize those results against the expected drug costs per hospitalization prevented.

## Methods

### Review of Literature and Estimations of Effect Size

To balance efficacy with potential toxicity, the outcome of interest we selected was all-cause hospitalization among outpatients. Where this was unavailable, we used COVID-19 related hospitalization (and indicated this). Of note, use of the later could underestimate toxicity, as hospitalizations due to drug side effects might be excluded. We used these results to calculate the relative risk for hospitalization with 95% confidence intervals.

Results for colchicine were taken from the only large outpatient randomized controlled trial, COLCORONA (4). For the inhaled corticosteroids, we used the results of our fixed-effects meta-analysis (5) of all available trials (18–21) with the caveat that the fixed-effects model may overestimate efficacy (I^2^ = 49.2%). For fluvoxamine, we obtained the number of all-cause hospitalizations in both arms directly from the authors of the three completed clinical trials (6,7,22). In the TOGETHER trial (7) the authors originally included ER visits of ≥6 hours as a proxy for hospitalization due to the prohibitive number of admissions in Brazil exhausting capacity. To be more conservative, we chose only to include patients who spent more than 24 hours in the emergency department as equivalent to being hospitalized. The trial results were combined using a fixed effects meta-analysis (I^2^ = 0.2%).

Results for outpatient anti-spike protein antibody randomized controlled trials were limited to most recent phase 3 studies as most used an integrated phase 1/2/3 design that led to multiple publications describing the same patients. We limited our analysis to the latest phase 3 studies for bamlanivimab/etesevimab (8), casirivimab/imdevimab (9) and sotrovimab (10). Bamlanivimab monotherapy was not included as it is no longer a recommended treatment. Of note, at the time of submission, bamlanivimab/etesevimab is not yet approved for use by Health Canada.

Results for outpatient antiviral therapies were limited to one conference abstract and an associated clinicaltrials.gov entry for remdesivir (11,12); an phase 3 trial manuscript and a press release for molnupiravir (13,15) (meta-analyzed using a fixed-effects model with I^2^ = 69% and the same caveat that this may overestimate efficacy); and a press release with results for two trials of nirmatrelvir with ritonavir (14) (I^2^=29.2%). Random-effects meta-analysis was conducted for a sensitivity analysis and are presented in the Supplement. Recognizing that the field moves very quickly, we have developed a webpage (https://read.idtrials.com/outptcovid) which will be updated monthly at least until the end of 2022 to contain the most up-to-date efficacy and Canadian cost data possible.

### Estimation of Canadian Costs

Where available, representative drug pricing was taken from the Quebec government formulary (23). We chose the price of budesonide for the inhaled corticosteroid analysis because it was the least expensive option for a 14 day supply at the trial doses used and because it was the first corticosteroid to demonstrate a reduction in hospitalizations (20). As specific formulary pricing was not available for all products due to a lack of transparency on the public costs, we used any publicly available data on Canadian purchasing and/or extrapolated cost data from other jurisdictions. For all anti-spike protein antibodies, assuming there would be competition, we used the price that Canada negotiated for bamlanivimab (24) converted to Canadian dollars. For the antiviral agents, we used the price estimated by the manufacturers for international high-income countries (25,26) and converted it to Canadian dollars. Prices for all injectable agents did not include the price of administration, which will vary greatly across the agents. Societal costs were also not factored in and beyond the scope of this analysis.

### Estimation of Events Prevented and Costs per Event Prevented

For each drug, we took the estimates of relative risk of hospitalization and the 95% confidence interval to generate the estimated absolute risk reduction (and confidence interval) assuming a moderate baseline risk of hospitalization of 5%. Sensitivity analyses were conducted for 2.5% (low) and 10% (high) risk of hospitalization. By dividing 100 by the absolute risk reduction (rounded up), we estimated the number needed to treat (NNT) to prevent one hospitalization with corresponding confidence intervals. We then multiplied the NNT by the drug costs per patient treated to arrive at the estimated drug cost to prevent one admission. For comparison, CIHI has estimated the cost of a COVID-19 hospitalization at $23000 (27).

## Results

The included studies are summarized in **Table** 1. The results of the analysis are presented in **Figure 1 and Table 2** with random effects meta-analysis presented in **Supplemental Figure 1**. The repurposed drugs colchicine, fluvoxamine, and the inhaled corticosteroids had smaller effect sizes and larger numbers needed to treat assuming a 5% hospitalization risk at 80 (48-667), 87 (50-2000), and 72 (45-400), respectively. By contrast, the antiviral and antibody therapies had larger effect sizes and smaller numbers needed to treat at 24 (22-29) for nirmatrelvir/ritonavir; 24 (21-49) for remdesivir; 52 (36-124) for molnupiravir; 25 (22-39) for sotrovimab, 29 (25-37) for casirivimab/imdevimab and 29 (24-50) for bamlanivimab/etesevimab.

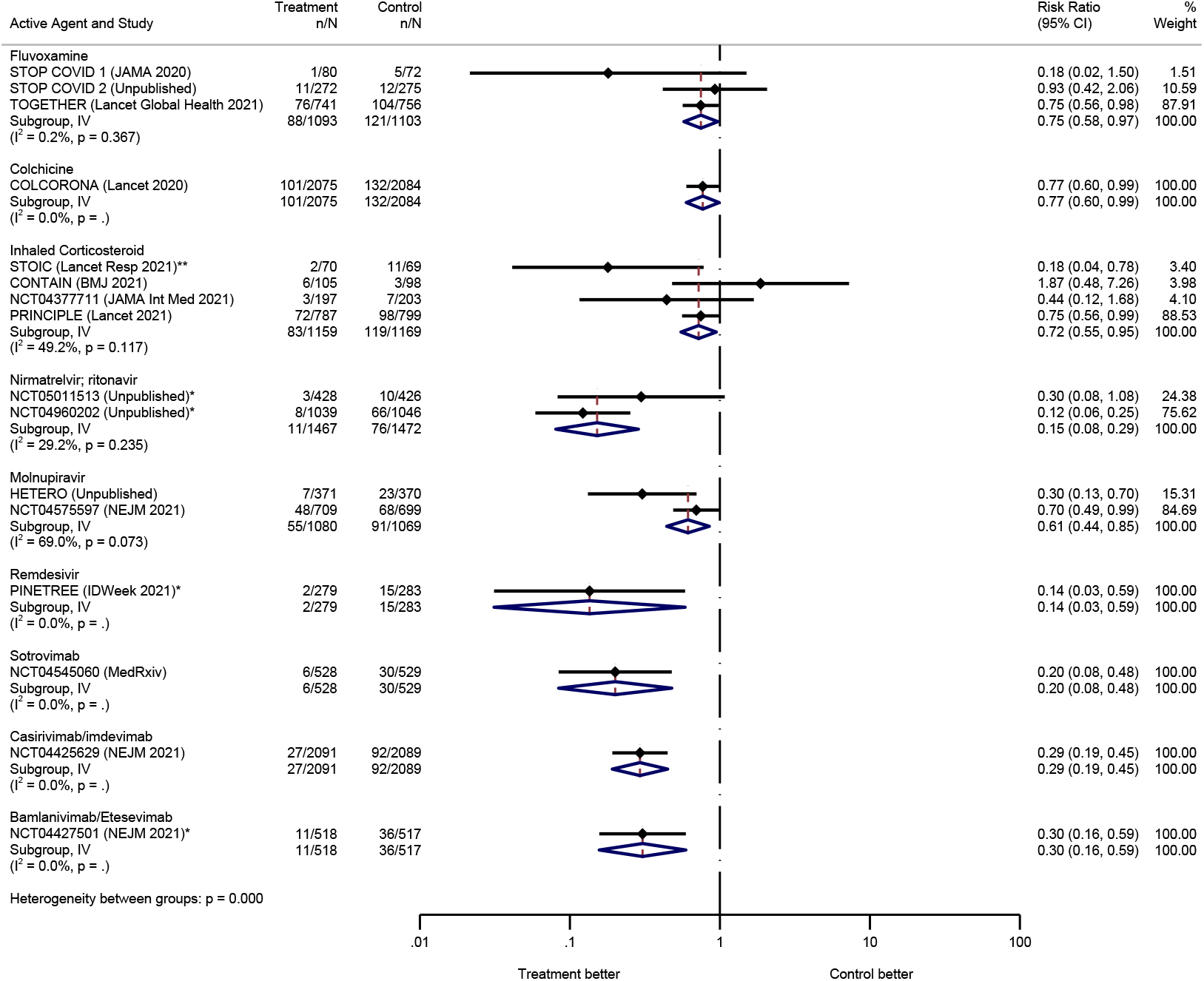

**Table 1.** Summary of included clinical trials.

| Study | Location | Original Primary Outcome | Inclusion Criteria | Demographics |
| --- | --- | --- | --- | --- |
| Fluvoxamine |  |  |  |  |
| Stop Covid 1 | USA | Clinical deterioration:<br>Hospitalization or new hypoxemia<br>within 15 days | Age $\geq 18$ unvaccinated<br>Positive test with:<br>$\leq 7$ days symptoms | Median age 46; 72% female; 70% white;<br>56% BMI $\geq 30$; 20% hypertension; 11% diabetes<br>Median 4 days of symptoms |
| Stop Covid 2 | USA and<br>Canada | Clinical deterioration:<br>Hospitalization or new hypoxemia<br>within 15 days | Age $\geq 30$ unvaccinated<br>Positive test with:<br>$\leq 6$ days symptoms<br>Criterion for high risk | Median age 47; 62% female; 73% white;<br>44% BMI $\geq 30$; 21% hypertension; 9% diabetes<br>Median 5 days of symptoms |
| Together | Brazil | ER visit $\geq 6$ hours or<br>hospitalization within 28 days | Age $\geq 18$ unvaccinated<br>Positive test with:<br>$\leq 7$ days symptoms<br>Criterion for high risk | Median age 50; 55% female; 96% mixed race;<br>51% BMI $\geq 30$; 13% hypertension; 16% diabetes<br>Mean 3.8 days of symptoms** |
| Colchicine |  |  |  |  |
| Colcorona | Multiple<br>countries<br>(majority<br>Canada) | COVID-19-related hospitalization<br>or death from any cause | Age $\geq 40$ unvaccinated<br>93% had positive test with:<br>Diagnosis within 24h<br>Criterion for high risk | Median age 53-54; 54% female; 93% white;<br>Mean BMI 30; 36% hypertension; 20% diabetes<br>Mean 5.3 days of symptoms |
| Inhaled Corticosteroids |  |  |  |  |
| STOIC* | UK | Covid-19 urgent visits | Age $\geq 18$ unvaccinated<br>94% had positive test with:<br>$\leq 7$ days of symptoms (at least one of<br>cough and fever or anosmia) | Mean age 45; 56% female; 93% white;<br>Mean BMI 26-27; N/A hypertension; 4% diabetes<br>Median 3 days of symptoms |
| CONTAIN | Canada | Resolution of cough, dyspnea, and<br>fever day 7 | Age $\geq 18$ unvaccinated<br>Positive test with:<br>$\leq 6$ days of symptoms (at least one of<br>fever, cough, or dyspnea) | Median age 35; 54% female; 61% white;<br>BMI not reported; 6% hypertension; 3% diabetes<br>Duration of symptoms not reported |
| Covis Pharma | USA | Time to symptom free | Age $\geq 12$ unvaccinated<br>Positive test with:<br>At least one of fever, cough, or dyspnea | Mean age 43; 55% female; 86% white;<br>Mean BMI 29.4; 22% hypertension; 8% diabetes<br>Duration of symptoms not reported |
| PRINCIPLE* | UK | COVID-19-related hospitalization or death from any cause | Age $\geq 65$ or $\geq 50$ with comorbidity<br>*14% vaccinated*<br>Positive test with ongoing symptoms of fever, new continuous cough, or change in smell or taste within 14 days | Mean age 64-65; 51% female; 93% white;<br>BMI not reported; 45% hypertension; 21% diabetes<br>Mean 6 days of symptoms |
| Nirmatrelvir/ritonavir |  |  |  |  |
| EPIC-HR (NCT04960202) | Multiple countries (USA 45%) | COVID-19-related hospitalization or death from any cause | Age $\geq 18$ unvaccinated<br>Positive test with:<br>$\leq 5$ days of symptoms (at least 1 at randomization)<br>Criterion for high risk | Not available at time of this analysis |
| EPIC-SR (NCT05011513) | Multiple countries | Time to Sustained Alleviation of All Targeted COVID-19 Signs/Symptoms | Age $\geq 18$ unvaccinated<br>Positive test with:<br>$\leq 5$ days symptoms<br>*Vaccinated if criterion for high risk* | Not available at time of this analysis |
| Molnupiravir |  |  |  |  |
| Hetero Pharma* | India | Hospitalization | Age $\geq 18$ and $\leq 60$ (vaccination unspecified)<br>Positive test with:<br>$\leq 5$ days of symptoms | Not available at time of this analysis |
| MOVE-Out (NCT04575597) | Multiple countries (majority Latin America) | All cause hospitalization or death (included ER visit $\geq 24$ hours) | Age $\geq 18$ unvaccinated<br>Positive test with:<br>$\leq 5$ days of symptoms (at least 1 at randomization)<br>Criterion for high risk | Median age 43; 51% female; 52% white (interim);<br>74% BMI $\geq 30$; N/A hypertension; 16% diabetes<br>48% had symptoms 3 days or less |
| Remdesivir |  |  |  |  |
| PINETREE | Multiple | COVID-19 related hospitalization | Age $\geq 18$ unvaccinated: | Mean age 50; 48% female; 80% white; |
|  | countries<br>(95% USA) | or all-cause death | Positive test with:<br>≤7 days of symptoms<br>Criterion for high risk | 56% BMI ≥30; 48% hypertension; 62% diabetes<br>Duration of symptoms not reported |
| Antibody therapies |  |  |  |  |
| Sotrovimab<br>(NCT04545060) | Multiple<br>countries<br>(92% USA) | Hospitalization for ≥24 hours or<br>death | Age ≥18 unvaccinated<br>Positive test with:<br>≤5 days of symptoms<br>Criterion for high risk | Interim analysis:<br>Median age 53; 54% female; 87% white;<br>63% BMI ≥30; N/A hypertension; 23% diabetes<br>59% had duration of symptoms ≤ 3 days |
| Casirivimab/<br>imdevimab<br>(NCT04425629) | USA and<br>Mexico | COVID-19-related hospitalization<br>or death from any cause | Age ≥18 unvaccinated<br>Positive test with:<br>≤7 days of symptoms<br>Criterion for high risk | Median age 48-50; 52% female; 84% white;<br>57% BMI ≥30; 36% hypertension; 15% diabetes;<br>Median duration of symptoms 3 days |
| Bamlanivimab/<br>etesevimab<br>(NCT04427501) | USA | COVID-19-related hospitalization<br>or death from any cause | Age ≥12 unvaccinated<br>Positive test with:<br>Symptoms<br>Criterion for high risk | Mean age 54; 52% female; 87% white<br>Mean BMI 34; 34% hypertension; 28% diabetes<br>Median duration of symptoms 4 days |
| ER=Emergency Room; *Open label; **Missing data on ~23% |  |  |  |  |

**Table 2.**
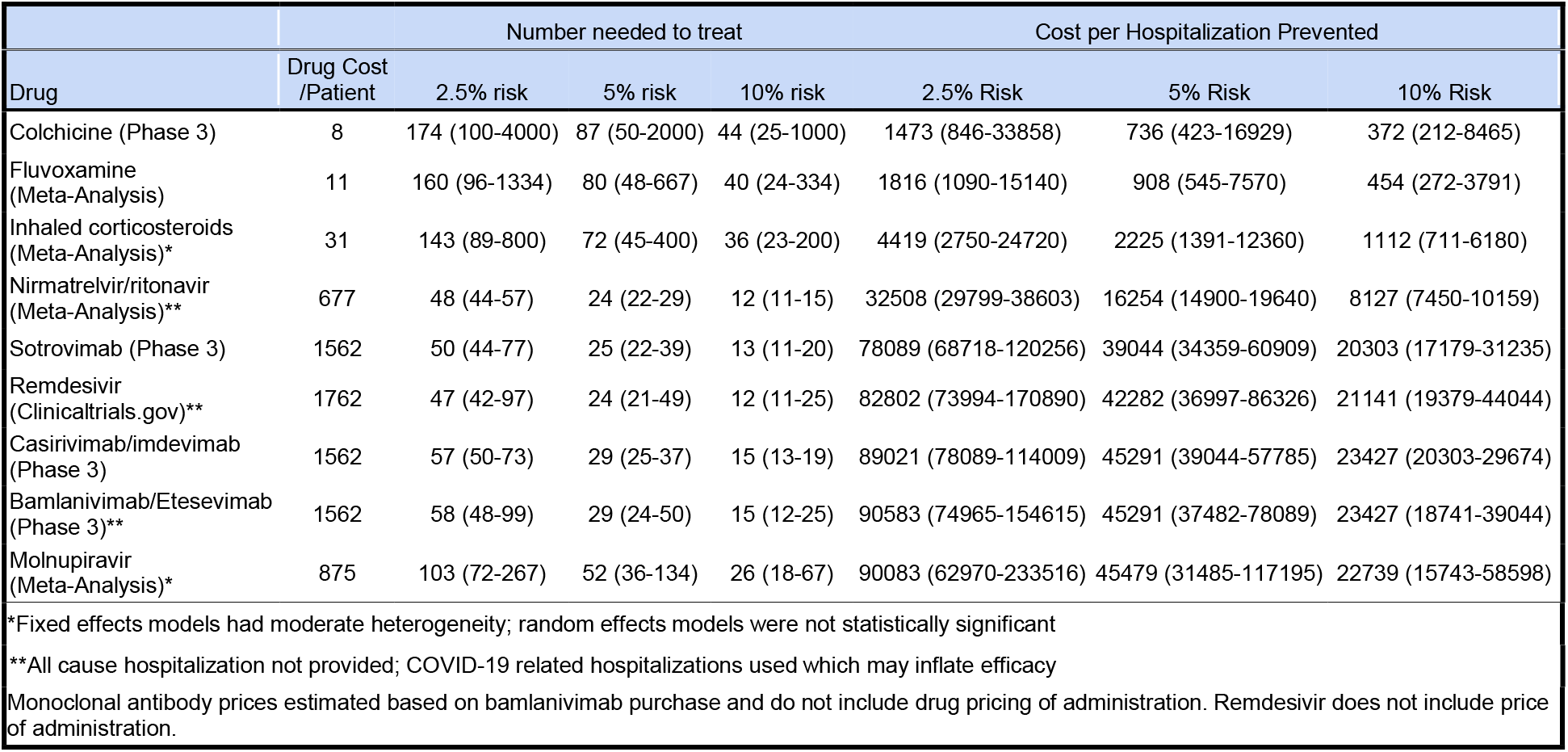
Number Needed to Treat and Costs per Hospitalization Prevented.

At a 5% risk of hospitalization, the corresponding drug costs per hospitalization prevented were: $736 ($423-$16929) for colchicine; $908 ($545-$7570) for fluvoxamine; $2225 ($1391-$12360) for inhaled corticosteroids; $16254 ($14900-$19640) for nirmatrelvir/ritonavir; $39044 ($34359-$60909) for sotrovimab; $42282 ($36997-$86326) for remdesivir; $45291 ($39044-$57785) for casirivimab/imdevimab; $45291 ($37482-$78089) for bamlanivimab/etesevimab and $45479 ($31485-$117195) for molnupiravir.

### Interpretation

Scientific advancement has been exponential during the pandemic. We have gone from the discovery of a new disease with no treatment in December 2019 to numerous effective vaccines coupled with a suite of proven therapies that prevent hospitalization and, by extension, presumably impact progression to death. Within these treatments, several are inexpensive and widely available worldwide, while others are expensive, limited in availability, and/or limited in accessibility. The purpose of this review was to contrast the efficacy of these therapies against a measure of cost, from the Canadian perspective. Evidence suggests that monoclonal antibody and antiviral therapies are more effective than repurposed drugs. While we found this efficacy comes at a price which often exceeds that of COVID-19 hospitalization, this was not a formal cost-effectiveness analysis and we did not factor in long term outcomes, societal costs, patient preferences nor costs of administration. The higher an individual’s baseline risk of deterioration, the greater the absolute benefit and less costly these options become. Furthermore, some of these therapies have already been purchased, which necessarily alters the dialogue. Essentially, at the right price or if prescribed to a high enough risk individual, every drug on this list has the potential to be cost saving to the system. Accurate Canadian models for prediction of hospitalization risk will be essential in contextualizing and maximizing the benefits of any therapy.

This analysis has several limitations. First, most of the agents studied have only had a single positive randomized controlled trial and several are pre-publication. Replication in science is important and while the pandemic necessitated speed which led to single trials, with many vaccinations and outpatient therapy options now available the safeguards of confirmatory trials are likely needed for reproducibility and generalizability. Second, for the antiviral agents, most data are currently limited to conference abstracts, trial registry, and press release. In normal times, peer reviewed results would be required; however, major policy decisions are being made based on industry public relations material, confidential submissions, and limited data (16,17,24) and our analysis can serve to inform those conversations. Third, our sensitivity analysis for inhaled corticosteroids and molnupiravir showed the benefits were not statistically significant in the random-effects meta-analysis and the results of our current analysis are predicated on additional trials confirming these drugs have benefit. Fourth, the efficacy of anti-spike protein antibodies require confirmation for new emerging variants, such as Omicron as there are suggestions that some therapies may no longer be as effective (28). Fifth, two of the trials have reported only interim results and the final efficacy may change as was seen with molnupiravir (15). Finally, very few patients in these clinical trials were vaccinated and the relative risk reduction for hospitalization may not be the same in vaccinated patients.

There is an ongoing need to identify effective treatments that can be administered early in the disease to prevent COVID-19 hospitalization and death and to make them available and accessible in all regions. While many Canadians are fortunate to have access to novel treatments, the number and location of available doses may wax and wane over time and access may be challenging in remote regions or in congregate care settings. Some degree of decision making will be required at the level of the individual clinician, hospitals, provinces, and the federal government to prioritize deployment of therapies and capacity building. This analysis provides one means of contextualizing those discussions.

## Supporting information

Supplemental Figure 1 - Random Effects Meta-Analysis

## Data Availability

All data produced in the present work are contained in the manuscript

## CRediT author statement

Conceptualization - TCL, EGM; Methodology - TCL, SAG, EGM; Validation - TCL; Formal Analysis - TCL; Investigation - All authors; Resources - TCL; Data Curation - TCL, EGM; Writing - Original Draft - TCL, EGM; Writing - Review and Editing - All authors; Visualization TCL, EGM

## Notes

Declarations: TCL and EGM receive research salary support from the Fonds de Recherche du Québec – Santé.

**Conflicts of Interest** TCL and EGM were co-investigators on several outpatient drug repurposing RCTs for COVID-19 (hydroxychloroquine, inhaled ciclesonide, and fluvoxamine).

### Competing Interest Statement

TCL and EGM receive research salary support from the Fonds de Recherche du Quebec - Sante.
TCL and EGM were co-investigators on several outpatient drug repurposing RCTs for COVID-19 (hydroxychloroquine, inhaled ciclesonide, and fluvoxamine).

### Funding Statement

This study did not receive any funding

