## Supplemental Figure 1 - Random Effects Meta-Analysis for "Outpatient therapies for COVID-19: How do we choose?"

| Active Agent and Study | Treatment n/N | Control n/N | Risk Ratio (95% CI) | % Weight |
|---|---|---|---|---|
| Fluvoxamine | | | | |
| STOP COVID 1 (JAMA 2020) | 1/80 | 5/72 | 0.18 (0.02, 1.50) | 1.51 |
| STOP COVID 2 (Unpublished) | 11/272 | 12/275 | 0.93 (0.42, 2.06) | 10.59 |
| TOGETHER (Lancet Global Health 2021) | 76/741 | 104/756 | 0.75 (0.56, 0.98) | 87.91 |
| Subgroup, REML | 88/1093 | 121/1103 | 0.75 (0.58, 0.97) | 100.00 |
| ($I^2$ = 0.2%, p = 0.367) | | | | |
| Colchicine | | | | |
| COLCORONA (Lancet 2020) | 101/2075 | 132/2084 | 0.77 (0.60, 0.99) | 100.00 |
| Subgroup, REML | 101/2075 | 132/2084 | 0.77 (0.60, 0.99) | 100.00 |
| ($I^2$ = 0.0%, p = .) | | | | |
| Inhaled Corticosteroid | | | | |
| STOIC (Lancet Resp 2021)** | 2/70 | 11/69 | 0.18 (0.04, 0.78) | 16.01 |
| CONTAIN (BMJ 2021) | 6/105 | 3/98 | 1.87 (0.48, 7.26) | 17.81 |
| NCT04377711 (JAMA Int Med 2021) | 3/197 | 7/203 | 0.44 (0.12, 1.68) | 18.16 |
| PRINCIPLE (Lancet 2021) | 72/787 | 98/799 | 0.75 (0.56, 0.99) | 48.02 |
| Subgroup, REML | 83/1159 | 119/1169 | 0.64 (0.31, 1.29) | 100.00 |
| ($I^2$ = 49.2%, p = 0.117) | | | | |
| Nirmatrelvir; ritonavir | | | | |
| NCT05011513 (Unpublished)* | 3/428 | 10/426 | 0.30 (0.08, 1.08) | 31.87 |
| NCT04960202 (Unpublished)* | 8/1039 | 66/1046 | 0.12 (0.06, 0.25) | 68.13 |
| Subgroup, REML | 11/1467 | 76/1472 | 0.16 (0.07, 0.37) | 100.00 |
| ($I^2$ = 29.2%, p = 0.235) | | | | |
| Molnupiravir | | | | |
| HETERO (Unpublished) | 7/371 | 23/370 | 0.30 (0.13, 0.70) | 39.23 |
| NCT04575597 (NEJM 2021) | 48/709 | 68/699 | 0.70 (0.49, 0.99) | 60.77 |
| Subgroup, REML | 55/1080 | 91/1069 | 0.50 (0.23, 1.11) | 100.00 |
| ($I^2$ = 69.0%, p = 0.073) | | | | |
| Remdesivir | | | | |
| PINETREE (IDWeek 2021)* | 2/279 | 15/283 | 0.14 (0.03, 0.59) | 100.00 |
| Subgroup, REML | 2/279 | 15/283 | 0.14 (0.03, 0.59) | 100.00 |
| ($I^2$ = 0.0%, p = .) | | | | |
| Sotrovimab | | | | |
| NCT04545060 (MedRxiv) | 6/528 | 30/529 | 0.20 (0.08, 0.48) | 100.00 |
| Subgroup, REML | 6/528 | 30/529 | 0.20 (0.08, 0.48) | 100.00 |
| ($I^2$ = 0.0%, p = .) | | | | |
| Casirivimab/imdevimab | | | | |
| NCT04425629 (NEJM 2021) | 27/2091 | 92/2089 | 0.29 (0.19, 0.45) | 100.00 |
| Subgroup, REML | 27/2091 | 92/2089 | 0.29 (0.19, 0.45) | 100.00 |
| ($I^2$ = 0.0%, p = .) | | | | |
| Bamlanivimab/Etesevimab | | | | |
| NCT04427501 (NEJM 2021)* | 11/518 | 36/517 | 0.30 (0.16, 0.59) | 100.00 |
| Subgroup, REML | 11/518 | 36/517 | 0.30 (0.16, 0.59) | 100.00 |
| ($I^2$ = 0.0%, p = .) | | | | |
| Heterogeneity between groups: p = 0.000 | | | | |

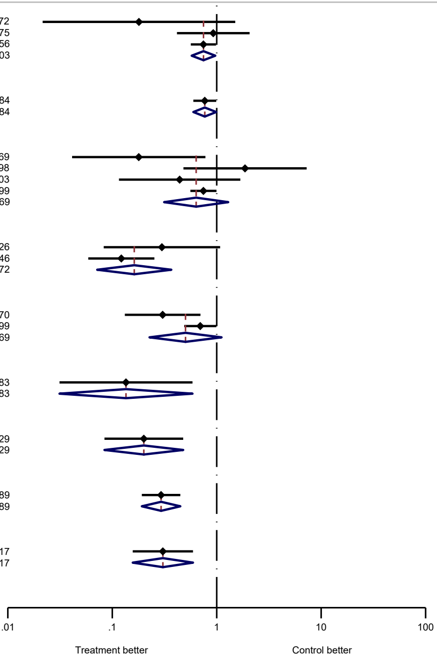


NOTE: Weights and between-subgroup heterogeneity test are from random-effects model
